# The impact of flavored e-cigarette bans on e-cigarette use in three US states

**DOI:** 10.1101/2023.05.19.23290249

**Authors:** Yong Yang, Eric N. Lindblom, Kenneth D. Ward, Ramzi G. Salloum

**Author notes:** Corresponding author: Yong Yang, Ph.D, Associate Professor, 203 Robison Hall, School of Public Health, University of Memphis, Memphis, TN, 38152.

## Abstract

**Introduction:** Beginning in 2019, several U.S. states implemented temporary or permanent bans on the sale of flavored e-cigarettes. This study examined the impact of flavor bans on adult e-cigarette use in Washington, New Jersey, and New York.

**Methods:** Adults who used e-cigarettes at least once a week before the flavor bans were recruited online. Respondents reported their e-cigarette use, primarily used flavor, and ways of obtaining ecigarettes before and after the bans. Descriptive statistics and multinomial logistic regression models were applied.

**Results:** After the ban, 8.1% of respondents (N=1624) quit using e-cigarettes, those primarily used banned menthol or other flavors declined from 74.4% to 50.8, those using tobacco-flavored declined from 20.1% to 15.6%, and those using non-flavored increased from 5.4% to 25.4%. More frequent e-cigarette use and smoking cigarettes were associated with being less likely to quit e-cigarettes and more likely to use banned flavors. Of those primarily using banned flavors, 45.1% obtained e-cigarettes from in-state stores, 31.2% from out-of-state stores, 32% from friends, family, or others, 25.5% from Internet/mail sellers, 5.2% from illegal sellers, 4.2% mixed flavored e-liquids themselves, and 6.9% stocked up on e-cigarettes before the ban.

**Conclusions:** Most respondents continued to use e-cigarettes with banned flavors post-ban. Compliance of local retailers with the ban was not high, and many respondents obtained banned-flavor e-cigarettes through legal channels. However, the significant increase in the use of non-flavored e-cigarettes post-ban suggests that these may serve as a viable alternative among those who used previously used banned or tobacco flavors.

**Implications:** This study examined the impact on adult e-cigarette users from the recent e-cigarette-only flavor bans in Washington State, New Jersey, and New York. We found that most respondents continued to use e-cigarettes with banned flavors post-ban and obtain banned-flavor e-cigarettes through legal channels. Our findings indicate that non-flavored e-cigarettes may serve as an acceptable alternative to both non-tobacco and tobacco-flavored e-cigarettes and state e-cigarette flavor bans are unlikely to prompt a significant number of adult e-cigarette users to replace their e-cigarette use with new or increased smoking. Enforcing compliance of retailers to the policy is crucial to control e-cigarette use.

## Introduction

The role of e-cigarette flavors, and restrictions on those flavors, in the initiation, use, and cessation of e-cigarettes and smoked tobacco products, including switching between combustible tobacco products and e-cigarettes or engaging in dual-use, is of enormous interest to the FDA and other policymakers (1). Assuming that e-cigarette use is a less-harmful alternative to smoking, flavored e-cigarettes could reduce health harms by attracting smokers and prompting them to switch to e-cigarettes (2-4) or by prompting youth who would otherwise become smokers to use e-cigarettes, instead. At the same time, however, flavors can increase initiation of e-cigarette use among youth and young adults who would not otherwise use any tobacco or nicotine product (5-8), and the initiation of e-cigarette use may serve as a precursor to regular cigarette smoking, causing additional new harms among those who otherwise would not smoke (9, 10).

In 2009, the U.S. Tobacco Control Act banned all characterizing flavors except menthol and tobacco in cigarettes. The rise in e-cigarette use, particularly among youth, and the outbreak of sudden lung injuries associated with vaping in 2019, prompted the FDA and several state and local governments to ban or consider banning all or some e-cigarettes, such as any with added flavors other than tobacco. At the beginning of 2020, the FDA initiated a temporary enforcement policy that banned the sale of all capsule-based e-cigarettes with flavors other than tobacco or menthol but did not apply the flavor restrictions to disposable or open-systems e-cigarettes (i.e., those with a chamber that users can fill with the e-liquid of their choice). Several states and local governments also took action to restrict flavored e-cigarettes. For example, bans on the sale of all e-cigarettes with added flavors other than tobacco were temporarily enforced in Montana (December 2019 to April 2020) and Washington (October 2019 to February 2020) and were permanently established in New Jersey (April 2020), New York (May 2020), and Rhode Island (March 2020).

Initial evidence suggested that the state and local flavor bans might not sharply reduce the availability or use of flavored tobacco products among residents because users could still obtain banned tobacco products through purchases in nearby jurisdictions with no bans, online, from non-complying in-state retailers, or from online sellers or other illegal sellers (11). At the same time, there have been concerns that e-cigarette flavor bans might prompt some smokers or former smokers using e-cigarettes to return to exclusive smoking. But the actual impact of flavor restrictions at the state or local level on adult and youth tobacco use patterns is unclear. To address the adult-side of this knowledge gap, this study examined the impact on adult e-cigarette users from the recent e-cigarette-only flavor bans in Washington State, New Jersey, and New York (Rhode Island and Montana were not included because of their relatively small populations).

## Methods

### Survey

Data were collected on Amazon Mechanical Turk (MTurk) (12), an efficient and cost-effective online crowdsourcing platform that has been used widely in tobacco studies (13-20). The survey was active in MTurk for two weeks in February 2020 for Washington and June 2020 for New Jersey and New York. Inclusion criteria were: 18 years of age or older; a current resident who has resided in the state during the past six months; regularly used e-cigarettes (at least once a week) for at least six months before the survey; and ≥90% approval rating from previous MTurk tasks. Eligible respondents were given access to the survey, hosted by Qualtrics (Provo, UT). The “Prevent Ballot Box Stuffing” option provided by Qualtrics was used to prevent respondents from taking the survey multiple times. To further increase the quality of the survey and prevent fake information, we used a zipcode double-checking mechanism (11). The Institutional Review Board at the University of Memphis approved this study.

### Primary measures

Respondents reported several features of their e-cigarette use for the past 30 days (for post-ban) and in the month before the state flavor ban (for pre-ban). For use frequency pre-ban, respondents reported whether they used e-cigarettes daily (every day or most days in a week) or weekly (at least once a week, but not most days). For post-ban, respondents reported whether they used e-cigarettes daily, weekly, less than weekly, or not at all. For both pre- and post-ban, respondents reported all e-cigarette flavors they used, the flavor that they primarily used, and the different ways they obtained e-cigarettes.

In addition to demographic information such as age, gender, race/ethnicity, educational attainment, and household income, we collected several variables that potentially impacted respondents’ responses to the ban, including how many years in total they had regularly used e-cigarettes, the extent to which they wanted to quit using e-cigarettes before the ban, their reasons for using e-cigarettes (i.e., for their flavors or to help in quitting smoking cigarette), and their cigarette smoking status. Respondents were also asked whether they were aware of the ban, supported the ban, and extent to which they perceived local retailers were compliant with the ban.

### Analysis

First, we depicted the characteristics of respondents from the three states with chi-square tests to evaluate demographic differences by state and the distribution of respondents by their tobacco and nicotine use status. Second, we reported the distribution of the primarily used e-cigarette flavor before and after the ban with paired t-tests to evaluate the differences. Third, a multinomial logistic regression (PROC LOGISTIC in SAS, version 9.4) was used to estimate the associations between demographics, e-cigarettes and smoking preference and use status, and the flavor that respondents primarily used after the ban. Fourth, we reported the distribution of ways of obtaining e-cigarettes before and after the flavor ban. Finally, the use of non-e-cigarettes tobacco products after the ban among different categories of respondents was assessed.

## Results

As shown in Table 1, the majority of the 1624 respondents were males (61.2%), young adults between 25 and 34 years old (56.9%), Whites (59.4%), and had at least a bachelor’s degree (66.8%). The majority of respondents had used e-cigarettes between two and five years (69.6%) and 79.8% had moderate or strong intentions to quit. The percent of respondents saying they used e-cigarettes because of the added flavors or for quitting smoking cigarettes were 54.4% and 36.6%, respectively. The majority of respondents currently smoked cigarettes either daily (30.9%) or weekly (44.4%). Most respondents had been aware of the ban before the survey (67.7%), with a plurality feeling neutral about the ban (36.1%) but more supporting the ban than opposing it (35.9% vs. 28.1%). Among respondents who knew about the ban before the survey, the perceived compliance of local retailers with the ban as lower than 50% was quite high (66.2%), and relatively few perceived that more than 75% of retailers were complying (14.8%). Respondents from New Jersey and New York were similar overall, and respondents from Washington were significantly different as they had a relatively lower proportion of males, Whites, with high educational attainment, and were less supportive of the ban but perceived higher compliance of local retailers with the ban compared with respondents from the other two states.

**Table 1.** Participants’ demographic characteristics, the status of e-cigarettes and tobacco use, and attitude/perception to the flavor ban (N=1624)

|  |  | All,<br>N=1624 | New Jersey,<br>N=600 | New York,<br>N=745 | Washington,<br>N=279 |
| --- | --- | --- | --- | --- | --- |
| Gender | Male | 61.2 | 63.3 | 61.7 | **54.8\*\*** |
|  | Female | 38.9 | 36.7 | 38.3 | **45.2\*\*** |
| Age, in years | 18-24 | 13.7 | 11.2 | 13.6 | **19.7\*\*** |
|  | 25-34 | 56.9 | 59.5 | 58.1 | **48.0\*\*** |
|  | 35-54 | 26.1 | 26.0 | 25.1 | **29.0\*\*** |
|  | 55 and above | 3.3 | 3.3 | 3.2 | **3.2\*\*** |
| Race /ethnicity | White | 59.4 | 53.5 | 55.6 | **64.9\*\*** |
|  | Black | 13.9 | 15.0 | 15.7 | **6.8\*\*** |
|  | Asian | 8.5 | 8.5 | 8.3 | **9.0\*\*** |
|  | Hispanic | 18.2 | 19.7 | 18.0 | **15.4\*\*** |
|  | Other/mixed race | 3.0 | 3.3 | 2.4 | **3.9\*\*** |
| Educational attainment | High school or less | 7.6 | 6.8 | 7.1 | **10.8\*\*** |
|  | Less than bachelor and more than high school | 25.6 | 22.8 | 23.6 | **36.6\*\*** |
|  | Bachelor or higher | 66.8 | 70.3 | 69.3 | **52.7\*\*** |
| Household income | Less than \$24,999 | 17.9 | 15.3 | **19.2\*** | **19.7\*** |
| | \$25,000 to \$49,999 | 32.3 | 34.7 | **30.7\*** | **31.2\*** |
| | \$50,000 to \$74,999 | 23.8 | 25.0 | **24.3\*** | **19.7\*** |
| | \$75,000 to \$99,999 | 14.2 | 13.2 | **14.2\*** | **16.1\*** |
| | \$100,000 or more | 12.0 | 11.8 | **11.5\*** | **13.3\*** |
| Smoking status (cigarettes or cigars) | Daily or most days of the week | 30.9 | 32.2 | 29.7 | 31.2 |
|  | Weekly smoker | 44.4 | 45.5 | 46.9 | **35.5\*\*** |
|  | Former smoker | 19.2 | 16.5 | 17.7 | **28.7\*\*** |
|  | Never smoker | 5.6 | 5.8 | 5.8 | **4.7\*** |
| History using e-cigarettes | <=1 year | 14.8 | 12.5 | **16.0\*** | **16.5\*\*** |
|  | 2-3 years | 44.5 | 44.5 | 42.8 | **49.1\*\*** |
|  | 4-5 years | 25.1 | 26.0 | 26.2 | **20.1\*\*** |
|  | >5 years | 15.6 | 17.0 | 15.0 | **14.3\*\*** |
| Reason for using e-cigarettes | It comes/came in flavors I like/liked | 54.4 | 52.8 | 53.8 | **59.1\*\*** |
|  | Using them helps people to quit smoking cigarettes | 36.6 | 37.3 | 33.6 | **43.0\*\*** |
| Frequency of e-cigarettes use before the ban | Used e-cigarettes daily | 44.5 | 41.5 | 41.2 | 59.5 |
|  | Used e-cigarette weekly | 55.5 | 58.5 | 58.8 | 40.5 |
| Intention to quit using e-cigarettes before the ban | Very much | 24.1 | 25.7 | 26.3 | **14.7\*\*** |
|  | Somewhat | 55.7 | 57.7 | 56.9 | **48.0\*\*** |
|  | Not all | 20.3 | 16.7 | 16.8 | **37.3\*\*** |
| Awareness of the ban when surveyed |  | 67.7 | 63.3 | **70.9\*\*** | **68.5\*** |
| Support for the ban when surveyed | Strongly supportive | 10.2 | 11.8 | 11.1 | **4.3\*\*** |
|  | Supportive | 25.7 | 26.5 | 28.5 | **16.5\*\*** |
|  | Neutral | 36.1 | 37.5 | 34.9 | **36.2\*\*** |
|  | Not supportive | 13.9 | 13.5 | 12.9 | **17.2\*\*** |
|  | Strongly not supportive | 14.2 | 10.7 | 12.6 | **25.8\*\*** |
| Perceived compliance of local retailers with the ban (among respondents aware of ban before the survey) | >75% | 14.8 | 11.3 | 13.1 | **26.7\*\*** |
|  | >50% - <=75% | 19.0 | 16.3 | 19.3 | **23.6\*\*** |
|  | >25% - <=50% | 34.8 | 36.1 | 35.6 | **29.8\*\*** |
|  | <=25% | 31.4 | 36.3 | 32.0 | **19.9\*\*** |

Note: boldface indicates the statistical significance of chi-tests, with the state of New Jersey as the reference, \* for P<0.05, and \*\* for P<0.01
<sup>1</sup> Any flavors other than tobacco and menthol, such as fruit, coffee, and candy

As shown in Table 2, in response to the flavor bans, 8.3% of respondents quit using e-cigarettes, the percentage of participants who primarily used menthol or who used other banned flavors decreased (from 19.6% to 15.7% and from 54.8% to 35.1%, respectively), and the portion primarily using non-flavored e-cigarettes increased (from 5.4% to 25.4%). However, the portion primarily using non-banned, tobacco-flavored e-cigarettes also decreased (from 20.1% to 15.6%). All pre-ban to post-ban changes in flavor use were statistically significant in paired t-test. Stratified analyses showed that respondents who primarily used non-TM flavors (i.e., any flavors other than tobacco and menthol, such as fruit, coffee, and candy) pre-ban were more likely to continue with the same primary flavor post-ban than respondents who primarily used menthol flavor (57.1% vs. 46.7%), but they were also more likely to quit using e-cigarettes (11.1% versus 5.6%). Compared with respondents who used e-cigarettes daily, respondents who used e-cigarettes weekly were more likely to decrease their use of menthol and non-TM flavors and were also more likely to quit e-cigarettes (12% versus 3.7%).

**Table 2.** Changes of the primarily used e-cigarette flavor before and after the ban

|  |  | Before or after the ban | Percent of participants primarily using each e-cigarette flavor |  |  |  |  |
| --- | --- | --- | --- | --- | --- | --- | --- |
|  |  |  | Tobacco | Menthol | Non-TM flavors <sup>1</sup> | Non-flavored | Quit e-cigarettes |
| All participants |  | Before | 20.1 | 19.6 | 54.8 | 5.4 | NA |
|  |  | After | 15.6 | 15.7 | 35.1 | 25.4 | 8.3 |
| Primary used flavor before the ban | Tobacco | After | 55.7 | 11.0 | 7.03 | 24.8 | 1.5 |
|  | Menthol | After | 8.8 | 46.7 | 10.7 | 28.2 | 5.6 |
|  | Non-TM flavors <sup>1</sup> | After | 4.2 | 7.8 | 57.1 | 19.9 | 11.1 |
|  | Non-flavored | After | 6.8 | 1.1 | 5.7 | 71.6 | 14.8 |
| Frequency of e-cigarette use before the ban | Daily | Before | 24.2 | 17.2 | 54.0 | 4.6 | NA |
|  |  | After | 18.6 | 16.1 | 40.2 | 21.5 | 3.7 |
|  | Weekly | Before | 16.9 | 21.6 | 55.4 | 6.1 | NA |
|  |  | After | 13.2 | 15.4 | 31.0 | 28.4 | 12.0 |
| Smoking status (cigarettes or cigars) | Daily or most days of the week | Before | 29.1 | 19.4 | 46.7 | 4.8 | NA |
|  |  | After | 21.2 | 17.8 | 33.5 | 24.4 | 3.2 |
|  | Weekly smoker | Before | 19.3 | 22.1 | 52.3 | 6.4 | NA |
|  |  | After | 15.7 | 17.1 | 31.6 | 27.9 | 7.8 |
|  | Former smoker | Before | 12.2 | 16.4 | 66.6 | 4.8 | NA |
|  |  | After | 10.0 | 10.0 | 43.1 | 23.5 | 13.5 |
|  | Never smoker | Before | 4.4 | 13.2 | 79.1 | 3.3 | NA |
|  |  | After | 3.3 | 13.2 | 44.0 | 16.5 | 23.1 |

Note: <sup>1</sup> Any flavor other than tobacco and menthol, such as fruit, coffee, and candy.
The flavors used most were significantly different before and after the ban according to the paired t-test.

As the stratified results by smoking status show, before the ban, the percent of e-cigarette users who primary used tobacco flavor was highest among respondents who were daily smokers (29.1%) and progressively declined along with the intensity of smoking, with former-smokers (12.2%) and never-smokers (4.4%) being least likely to primarily use tobacco-flavored e-cigarettes. The use of menthol flavor was also higher among smokers than former and never smokers. The percent of non-TM flavors had a reverse pattern, being lowest among daily smokers (46.7%), higher progressively along with those smoked weekly or less, former smokers, and highest among never smokers (79.1%). After the ban, the percentages of e-cigarette users who primarily used tobacco, menthol, and non-TM flavors decreased generally except the menthol share among never smokers did not change. After the ban, the percentage of those who quit using e-cigarettes was highest among never smokers (23.1%) and former smokers (13.5%) compared with 3.2% and 7.8% among those who smoked daily and weekly, respectively.

As shown in Table 3, after the ban younger age groups were more likely to use non-TM flavors, those with higher education were more likely to continue using banned flavors, and household income had little impact. Compared with other race/ethnicity groups, Hispanics were the least likely to quit and Blacks were the most likely to continue using banned flavors after the ban. Respondents were both less likely to quit e-cigarette use and more likely to continue using banned flavors if they had used e-cigarettes for a greater amount of time or had weaker intentions to quit before the ban. Respondents who used e-cigarettes because of the flavor were more likely to continue using banned flavors (statistically significant for non-TM flavors but not for menthol). Those primarily using non-TM flavors before the ban were most likely to quit using e-cigarettes and more likely to continue the same flavor afterward. Those primarily using menthol before the ban were both less likely to quit and less likely to continue the same flavor after the ban than those primarily using non-TM flavors before the ban. Those primarily using non-flavored e-cigarettes before the ban were more likely to quit than those primarily used tobacco flavor before the ban. People who used e-cigarettes daily before the ban were less likely to quit and more likely to keep using non-TM flavors. Never smokers and former smokers were more likely to quit e-cigarettes. While there were no significant differences in quitting all e-cigarette use after each of the three state bans, e-cigarette users in New York were less likely to use banned flavors after the ban than those in New Jersey.

**Table 3.** Multinomial logistic regression results of the flavor that respondents primarily used after the ban, using respondents who used tobacco flavor or non-favored e-cigarettes primarily as the reference group

|  |  | Quit (N=135, 8.3%) | Use menthol flavor (N=255, 15.7%) | Use non-TM flavors<sup>5</sup> (N=570, 35.1%) |
| --- | --- | --- | --- | --- |
| Gender | Male (ref) | 1 | 1 | 1 |
|  | Female | 0.68(0.44,1.04) | 1.24(0.89,1.74) | 1.15(0.87,1.53) |
| Age, in years | 18-24 (ref) | 1 | 1 | 1 |
|  | 25-34 | 0.82(0.44,1.52) | 0.81(0.45,1.45) | **0.62(0.40,0.95)\*** |
|  | 35-54 | 1.13(0.57,2.24) | 0.68(0.36,1.28) | **0.57(0.35,0.92)\*** |
|  | 55 and above | 1.04(0.33,3.22) | 0.38(0.12,1.18) | **0.32(0.13,0.79)\*** |
| Race/ethnicity | White (ref) | 1 | 1 | 1 |
|  | Black | 0.69(0.36,1.33) | 1.59(0.97,2.61) | **1.59(1.05,2.4)\*** |
|  | Asian | 0.77(0.39,1.51) | 0.71(0.36,1.38) | 1.08(0.66,1.76) |
|  | Hispanic | **0.3(0.13,0.66)\*\*** | 1.11(0.72,1.71) | 0.78(0.52,1.17) |
|  | Other/mixed race | 0.71(0.19,2.72) | 1.4(0.52,3.76) | 1.75(0.76,4.03) |
| <sup>1</sup> Educational attainment |  | 0.93(0.66,1.31) | 1.33(0.99,1.8) | **1.29(1.01,1.64)\*** |
| <sup>2</sup> Household income |  | 1(0.85,1.18) | 1.04(0.9,1.19) | 0.98(0.88,1.1) |
| <sup>3</sup> History using e-cigarette |  | **0.51(0.39,0.67)\*\*** | **1.35(1.13,1.62)\*\*** | **1.18(1.01,1.38)\*** |
| <sup>4</sup> Intention to quit using e-cigarettes before the ban |  | **0.57(0.41,0.79)\*\*** | 0.91(0.7,1.19) | **1.32(1.07,1.64)\*** |
| Using e-cigarettes because of a preferred flavor |  | 1.28(0.83,1.96) | 1.34(0.96,1.88) | **1.42(1.07,1.88)\*** |
| Using e-cigarettes to help quit smoking cigarette |  | 0.8(0.5,1.28) | 0.77(0.54,1.1) | 0.99(0.74,1.33) |
| Primarily used flavor before the ban | Tobacco (ref) | 1 | 1 | 1 |
|  | Menthol | **4.82(1.7,13.68)\*\*** | **11.39(7.17,18.09)\*\*** | **3.26(1.81,5.89)\*\*** |
|  | <sup>5</sup> Non-TM flavors | **12.58(4.89,32.39)\*\*** | **2.69(1.67,4.34)\*\*** | **24.45(15.18,39.37)\*\*** |
|  | Non-flavored | **5.38(1.75,16.56)\*\*** | **0.13(0.02,0.98)\*** | 0.81(0.29,2.25) |
| Frequency of e-cigarette use before the ban | Daily | 1 | 1 | 1 |
|  | Weekly | **2.15(1.26,3.67)\*\*** | 0.75(0.52,1.08) | **0.61(0.45,0.84)\*\*** |
| Smoking status (cigarettes or cigars) | Daily and most day in a week | 1 | 1 | 1 |
|  | Weekly | 1.28(0.66,2.46) | 1.11(0.74,1.67) | 1.08(0.76,1.55) |
|  | Former smoker | **3.22(1.62,6.39)\*\*** | 0.82(0.48,1.41) | 1.35(0.89,2.03) |
|  | Never smoker | **5.44(2.18,13.58)\*\*** | 1.81(0.75,4.35) | 1.53(0.77,3.05) |
| State | New Jersey (ref) | 1 | 1 | 1 |
|  | New York | 0.78(0.49,1.23) | **0.56(0.39,0.79)\*\*** | **0.66(0.49,0.89)\*\*** |
|  | Washington | 1.05(0.56,1.97) | 0.74(0.44,1.23) | 0.75(0.5,1.13) |

Notes: boldface indicates statistical significance, with \* for p<0.05, and \*\* for p<0.01
<sup>1</sup> Educational attainment categorized into three levels: 1. high school or less; 2. less than bachelor and more than high school; and 3. bachelor or more
<sup>2</sup> Household income was categorized into five levels: 1. Less than \$24,999; 2. \$25,000 to \$49,999; 3. \$50,000 to \$74,999; 4. \$75,000 to \$99,999; and 5. \$100,000 or more
<sup>3</sup> History using e-cigarettes was categorized into four levels: 1. <=1 year; 2. 2 and 3 years; 3. 4 and 5 years; and 4. >5 years
<sup>4</sup> Intention to quit using e-cigarettes before the ban was categorized into three levels: 1. Very much; 2. Somehow; and 3. Not all
<sup>5</sup> Any flavor other than tobacco and menthol, such as fruit, coffee, and candy.

As Table 4 shows, the percentage of respondents obtaining e-cigarettes from Internet/mail sources or from friends, family members, or other persons increased post-ban, compared with the portion of all respondents making such purchases pre-ban, not only among those who primarily used a banned flavor but also among those who primarily used non-banned tobacco flavor. After the ban, the Internet was used by about one-quarter of the respondents and about one-third of respondents obtained e-cigarettes from friends and other persons. While the percentages from vape shop/lounge purchases largely remained the same, the percentages obtaining e-cigarettes from tobacco specialty stores, grocery stores, and other general retailers decreased. Overall, after the ban, in-state stores remained the most important ways of obtaining e-cigarettes (41.8-45.1%) but purchases from stores out of state were also substantial (23.7-31.2%). Compared with those who primarily used tobacco-flavored e-cigarettes, respondents who primarily used banned flavors after the bans were more likely to engage in “unusual” ways of obtaining their e-cigarettes, such as from illegal sellers, mixing flavored e-liquids on their own, or by stocking up before the ban. Among respondents who continued to primarily use banned flavors post-ban, the greater they perceived the level of local retailer compliance with the ban, the more likely they were to purchase e-cigarettes via the Internet/mail and to have stocked up on e-cigarettes before the ban, and the less likely they were to purchase from either in-state or out-of-state stores or from vape shop/lounges, tobacco specialty stores, or grocery stores or other general retailers. Respondents who perceived the highest levels of retailer compliance (>75%) were also the least likely to obtain e-cigarettes from friends, family members, or other persons.

**Table 4.** Ways of obtaining e-cigarettes before and after the ban, percent (%)

|  | Before the ban (N=1624) | After the ban, among those who used tobacco flavor most (N=283) | After the ban, among those who used banned flavors e-cigarettes most (N=825) |  |  |  |  |  |  |  |
| --- | --- | --- | --- | --- | --- | --- | --- | --- | --- | --- |
|  |  |  | All (N=825) | By perceived compliance of local retailers with the ban among those who knew the ban (N=558) |  |  |  | By state (N=825) |  |  |
|  |  |  |  | >75%, N=73 | >50% - <=75%, N=106 | >25% - <=50%, N=186 | <=25%, N=193 | New Jersey, N=328 | New York, N=345 | Washington, N=152 |
| Internet/mail | 20.6 | 25.3 | 25.5 | 37.0 | 34.0 | 19.4 | 14.5 | 25.0 | 23.5 | 30.9 |
| Friends, family members, or other persons | 30.4 | 37.5 | 32.0 | 11.0 | 33.0 | 35.0 | 33.2 | 35.1 | 34.5 | 19.7 |
| Vape shop/lounge | 42.5 | 39.9 | 45.9 | 39.7 | 41.5 | 44.6 | 52.9 | 44.2 | 47.8 | 45.4 |
| Tobacco specialty stores | 27.0 | 17.4 | 21.2 | 12.3 | 19.8 | 20.4 | 29.0 | 22.3 | 23.2 | 14.5 |
| Grocery store, or other general retailers | 17.2 | 10.3 | 15.3 | 12.3 | 14.2 | 11.8 | 18.1 | 14.0 | 18.0 | 11.8 |
| Illegal sellers |  | 4.2 | 5.2 | 1.4 | 8.5 | 3.2 | 8.8 | 4.9 | 6.7 | 2.6 |
| Mixed flavored e-liquid by self |  | 1.4 | 4.2 | 4.1 | 2.8 | 2.7 | 8.8 | 4.0 | 4.63 | 4.0 |
| Stocked before the ban |  | 2.8 | 6.9 | 17.8 | 13.2 | 3.8 | 5.2 | 6.1 | 7.0 | 8.6 |
| Any stores within the State |  | 41.8 | 45.1 | 32.9 | 44.3 | 43.0 | 50.8 | 40.9 | 51.3 | 40.1 |
| Any stores out of the State |  | 23.7 | 31.2 | 30.1 | 24.5 | 29.0 | 41.5 | 35.1 | 29.0 | 27.6 |

Respondents who perceived the lowest levels of local retailers’ compliance (<25%) were more likely than those perceiving higher compliance to obtain e-cigarettes from illegal sellers (8.8%) and to mixed flavored e-liquid by themselves (8.8%). Respondents who primarily used banned flavors post-ban from Washington state were more likely to obtain e-cigarettes from the Internet and less likely to obtain from friends, family, or other persons compared with those from two other states who mostly used banned flavors.

## Discussion

A major concern regarding bans of flavored e-cigarette flavors (especially with no concurrent ban of flavored smoked tobacco products) is that they might prompt many smokers who switched to e-cigarettes to reduce or quit tobacco products to return to or increase their use of tobacco, or might prompt never-smoking users of flavored e-cigarettes to switch to flavored smoked tobacco products. Our findings suggest that these concerns may not be warranted, as only 8.3% of adult e-cigarette users quit using e-cigarettes post-ban, and being a nonsmoker or former smoker, rather than a smoker, was associated with a higher likelihood of quitting e-cigarette use.

As expected, quitting e-cigarettes was also associated with lower frequency of e-cigarette use and higher intentions to quit pre-ban. We also found that Hispanics were less likely to quit e-cigarette use in response to a state flavor ban than other ethnic groups, which is consistent with findings that racial/ethnic minorities are less likely to quit smoking successfully than non-Hispanic Whites (21, 22). As expected, respondents who primarily used banned-flavor e-cigarettes before the ban were more likely to quit than those who used tobacco flavor e-cigarettes, but it is noteworthy that respondents who primarily used non-flavored e-cigarettes were even more likely to quit. Besides factors such as quit intention, history of using e-cigarettes, and e-cigarette and cigarette use status that were already adjusted for in the regression analysis, there may be some characteristics of the group who used non-flavored e-cigarettes that warrant further examination.

Rather than quit, more than half of e-cigarette users simply continued using banned flavors despite the state bans, with many continuing to purchase from non-complying in-state sellers or other illegal sources and many purchasing from out-of-state sellers not subject to the bans. With stricter enforcement— and the new federal restrictions on e-cigarette sales via Internat/mail (23) — it is possible that more of these users would have quit e-cigarette use, with some possibly increasing their smoking, especially if menthol cigarettes or flavored cigars remained readily available. However, more than 40% of continuing e-cigarette users primarily used non-banned tobacco or non-flavored e-cigarettes post-ban, most likely because they did not feel comfortable purchasing from non-complying or otherwise illegal or inconvenient sellers or were unable to do so and were willing to accept tobacco-flavored or, even more, non-flavored e-cigarettes as suitable legal options. Accordingly, it is quite possible that most of those users who continued using banned-flavor e-cigarettes post-ban would have behaved similarly and switched to tobacco or non-flavored versions if they were unable to obtain e-cigarettes with banned flavors. The sharp increase in primary use of non-flavored e-cigarettes among all types of pre-ban e-cigarette users supports this conclusion. Moreover, the fact that many primary users of non-banned tobacco-flavored e-cigarettes switched to non-flavored e-cigarettes post-ban indicates that they can serve as a suitable alternative for primary users of tobacco-flavored e-cigarettes who want to move on to a non-tobacco version when all non-tobacco added flavors are no longer readily available.

It is likely, however, that many users who desire to do so would still be able to obtain banned-flavor e-cigarettes despite state bans, even with much stronger state enforcement and the new federal restrictions on Internet/mail sales (23), by purchasing from any remaining non-complying in-state retailers, from illegal in-state bootleg sellers, or from sellers in nearby states or Tribal lands not affected by the state bans, or by mixing their own e-cigarette flavors. While this study found quite low levels of buying from illegal sellers (5.2%) or mixing one’s own e-cigarette flavors (4.2%) among those primarily using banned flavors post-ban, buying from out-of-state sellers was much more prevalent (31.2%), and these methods of obtaining banned flavors could increase if other methods were more effectively restricted. Moreover, to the extent that those e-cigarette users who want to continue using banned flavors post-ban were able to do so, that would reduce the likelihood that they would quit e-cigarettes or turn to increased smoking, instead. In this study, for example, many of those perceiving high levels of in-state retailer compliance still reported considerable use of e-cigarettes with banned flavors.

Respondents who perceived a lower level of local retailers’ compliance to the ban were more likely to obtain e-cigarettes from illegal sellers and mixed flavored e-liquids on their own, compared with those who perceived a higher level of local retailer compliance. A possible explanation is the neighborhood effect, that is, in neighborhoods where law enforcement is relatively weak, local retailers are less likely to follow the ban and illegal sellers are more active. Our additional analyses (not reported here) showed that a significant difference in compliance exists both between states and between neighborhoods. Respondents living in Washington perceived higher compliance than respondents living in New York and New Jersey, and respondents living in suburban areas perceived higher retailer compliance than respondents living in urban and rural areas. Further studies will be needed to examine the variation of compliance across different geographic areas.

The large increase in the percent of respondents using non-flavored instead of flavored e-cigarettes after the bans could produce public health gains, as added flavors can increase the toxicity of inhaled e-cigarette aerosols (24, 25). Both for that reason and to ensure that non-flavored e-cigarettes will remain available as an acceptable alternative that smokers primarily using tobacco or other flavored e-cigarettes can move to after a flavor ban instead of increasing their smoking, policymakers should ensure that non-flavored e-cigarettes remain legally available. In addition, further research is needed to explore whether allowing only non-flavored e-cigarettes might be more beneficial for public health than also allowing tobacco-flavored versions. While state and local governments, and the FDA to some extent, have considered making tobacco flavor the default for e-cigarettes or the flavor that should most certainly be allowed despite flavor bans, it could very well be that the optimal regulatory approach is to allow only non-flavored e-cigarettes to minimize unnecessary additives and related actual or potential toxicity increases. This is especially important if other measures are implemented to push smokers away from smoked tobacco products or to attract smokers to e-cigarettes, making using flavors to attract smokers less necessary or unnecessary at all.

The study had several limitations. First, we did not collect detailed information on respondents’ use of other types of tobacco products both before and after the ban. Evidence shows that a ban on flavored tobacco products may prompt users to switch to other tobacco products (11, 26-28), thus besides the change in e-cigarette use, future research should more thoroughly examine how the use of other tobacco/nicotine products is impacted by the ban policy, to obtain a more complete picture. Second, the surveys were conducted during the first half of 2020, and respondents’ e-cigarette use patterns may have been influenced by the emerging COVID-19 pandemic. In addition, a new federal law restricting and regulating e-cigarette Internet/mail sales and related deliveries has been implemented since the study period, which should make it more difficult for users to obtain state-banned e-cigarettes through such sales or otherwise obtain e-cigarettes through the Internet/mail purchases. Third, this was a cross-sectional study with a non-representative enient sample and respondents may not have recalled their e-cigarette use before the ban precisely. Fourth, some measures such as the perceived compliance may be biased by the respondents’ e-cigarette use, and objective measures are needed to examine retailers’ compliance.

Despite these limitations, our findings indicate that non-flavored e-cigarettes may serve as an acceptable alternative to both non-tobacco and tobacco-flavored e-cigarettes and state e-cigarette flavor bans are unlikely to prompt a significant number of adult e-cigarette users to replace their e-cigarette use with new or increased smoking. A complete flavor ban (i.e., ban all added flavors) and enforcing compliance of retailers to the policy is crucial to control e-cigarette use.

## Data Availability

All data produced in the present study are available upon reasonable request to the authors

